# Relationship between temporal projections and cognition in patients with putaminal hemorrhage

**DOI:** 10.1101/2024.02.13.24302802

**Authors:** Sung Ho Jang, Hyeok Gyu Kwon

**Affiliations:** Department of Physical Medicine and Rehabilitation, College of Medicine, Yeungnam University; Department of Physical Therapy, College of Health Science, Eulji University

**Keywords:** Cognition, Diffusion tensor tractography, Putaminal hemorrhage, Temporal projection, Uncinate fasciculus

## Abstract

**Background:** The cognitive function is a complex mental process involving the temporal projections (cingulum, superior longitudinal fasciculus[SLF], inferior longitudinal fasciculus[ILF], and uncinate fasciculus[UF]). Therefore, a brain injury involving the temporal projections results in cognitive impairment. However, few studies have reported the pathophysiological mechanisms associated with cognitive impairment in patients with putaminal hemorrhage(PH). In this study, using diffusion tensor tractography(DTT), the relationship between the temporal projections and cognition was analyzed in patients with PH.

**Methods:** 25 patients with PH were recruited for this study. A Mini-Mental State Examination(MMSE) was conducted twice to evaluate their cognitive function(within one month and six months after onset). The temporal projections(cingulum, SLF, ILF, and UF) were reconstructed and the values of DTT parameters were measured. Pearson correlation and multiple regression analyses were also performed to determine the relationship between the DTT parameters of the temporal projections and the MMSE.

**Results:** An analysis of the correlations showed that the fractional anisotropy and fiber number ratio of the UF were correlated with the MMSE performed within one month and six months after onset. In addition, the fiber number of the UF on the MMSE at six months from the onset had an explanatory power of 33% and showed the 0.064 for unstandardized coefficients.

**Conclusions:** Using DTT, this study demonstrated that among the temporal projections, the state of the UF in patients with PH could be the most significant indicator of cognitive function. Our results suggested that evaluation of the UF using DTT could be useful in predicting the cognitive function following PH.

## Introduction

Spontaneous intracerebral hemorrhage accounts for 10-20% of cerebral strokes of which putaminal hemorrhage (PH) is the most common.^1–3^ Because of the location, the clinical presentations of PH not only include motor weakness and somatosensory disorder, but also cognitive impairment.^4–6^ Cognitive functions, including perception, attention, memory, and language processing are associated with various neural projections.^7,8^ In particular, temporal lobe white matter projections including the cingulum, superior longitudinal fasciculus (SLF), inferior longitudinal fasciculus (ILF), and uncinate fasciculus (UF) are mainly involved with cognitive function by connecting different brain regions:^9–17^ 1) The cingulum connects the orbitofrontal, parietal, and medial temporal lobes with the limbic system and is involved in attention, memory, learning, motivation, and emotion.^9–12^ 2) The SLF connects the frontal, parietal, and temporal lobes and is involved in motor control, working memory, language processing, and visual recognition.^13,14^ 3) The ILF connects the occipital and temporal lobes and is involved in visual and language processing.^15^ 4) The UF connects the frontal and temporal lobe and is involved in memory, emotion, and language processing.^16,17^ Therefore, any injury to the temporal projections affects cognitive function.^9–17^ As cognition is affected through interaction of each temporal projection, it is important to know which neural projections influence cognition and to what extent. However, relatively few studies have been reported on the pathophysiological mechanisms associated with cognitive impairment in patients with PH.^4,18–21^

Diffusion tensor tractography (DTT) derived from diffusion tensor imaging (DTI) is one of the non-invasive methods for investigating the human brain particularly its white matter. It offers a 3D visualization of specific neural projections and can quantify the integrity of the neural projections using fractional anisotropy (FA), apparent diffusion coefficient (ADC), and fiber number (FN).^22^ Hence, many studies have used DTT to evaluate the state of the temporal projections in patients with brain injury.^4,16–18,23–25^ However, there has been no study to date that has sought to understand which temporal projection at the early stage contributes more to the cognitive function at the chronic stage following PH.

In the current study, using DTT, we investigated the relationship between the temporal projections and cognition in patients with PH.

## Methods

### Subjects

A total of 25 patients (male: 17, female: 8, mean age: 51.9±14.2 years, range: 31∼ 74 years) who had been admitted to the rehabilitation department of a university hospital were recruited for this study. Inclusion criteria for patients were as follows: (1) First ever stroke, (2) A hematoma located primarily in the lentiform nucleus of the basal ganglia, (3) A DTI scan performed within one month of onset, and 4) Absence of hydrocephalus, subarachnoid hemorrhage, or intraventricular hemorrhage. This study was conducted retrospectively, and the study protocol was approved by the Institutional Review Board of the university hospital.

### Clinical evaluation

The cognitive function of the patients was evaluated using the Mini-Mental State Examination (MMSE) performed concurrently with DTI scanning (within one month from onset) and at six months after onset. The reliability and validity of the MMSE have been well established.^26^

### Diffusion tensor tractography

A 6-channel head coil on a 1.5T Philips Gyroscan Intera (Philips, Ltd., Best, The Netherlands) with single-shot echo-planar imaging was used for the acquisition of DTI data at a mean of 22.5±15.8 days. For each of the 32 non-collinear diffusion-sensitizing gradients, 70 contiguous slices parallel to the anterior commissure-posterior commissure line were acquired. The imaging parameters were as follows: Acquisition matrix = 96 × 96; reconstructed to matrix = 192 × 192; field of view = 240 × 240 mm^2^; TR = 10,726 ms; TE = 76 ms; parallel imaging reduction factor (SENSE factor) = 2; EPI factor = 49; *b* = 1000 s/mm^2^; NEX = 1; and a slice thickness of 2.5 mm. Removal of eddy current-induced image distortions was performed at the Functional Magnetic Resonance Imaging of the Brain (FMRIB) software library (FSL;www.fmrib.ox.ac.uk/fsl, Oxford, UK) using affine multi-scale two-dimensional registration. Reconstruction of the temporal projections was performed using DTI-Studio software (CMRM, Johns Hopkins Medical Institute, Baltimore, MD, USA).^27^ For the study, four temporal projections were selected (cingulum, SLF, ILF, and UF). For the cingulum, two regions of interest (ROIs) were drawn at the anterior and posterior green portion of the cingulum area on the color map of the coronal image.^28^ For the SLF, a triangular shape and green portion just lateral to the corticospinal tract near the anterior (first ROI) and posterior (second ROI) horn of the lateral ventricle on the color map of the coronal image were drawn.^29^ For the ILF, the first ROI was placed on the surrounding white matter of the anterior temporal lobe on the axial slice, and the second ROI on the surrounding white matter of the occipital lobe.^30^ For the UF, the first ROI was placed at the frontal part of the UF portion in the coronal image through the genu of the corpus callosum, just anterior to the anterior horn of the lateral ventricle, and the second ROI on the white matter in the coronal plane at the most anterior part of the temporal stem.^31^ Fiber tracking was started at any seed voxel with fractional anisotropy (FA) > 0.2 and ended at a voxel with a FA of < 0.2 and a tract turning-angle of < 60 degrees. The values of FA, ADC, and FN for the temporal projections were measured.

### Statistical analysis

The SPSS software (SPSS for Windows, Version 15.0; SPSS Inc., Chicago, USA) was used for statistical analysis. An independent t-test was used to compare the FA, ADC, and FN values between the affected and unaffected sides. Using the Pearson correlation, the MMSE score within one month and at six months of onset was used to determine the correlation with FA, ADC, and FN of the ratio (affected/unaffected) of temporal projections (cingulum, SLF, ILF, and UF). The correlation coefficient (*r*) indicates the relative strength (0.1–0.3: weak, 0.3–0.5: moderate, > 0.5: strong) and direction (+, −) of the linear relationship between two values.^32^ The contributions of the FA, ADC, and FN of the ratio (affected/unaffected) of temporal projections to the MMSE performed six months after onset were evaluated, using multiple regression. The null hypothesis of no difference was rejected if the p-values were less than 0.05.

## Results

A summary of the results for the DTT parameters for the temporal projections is shown in Table 2. A comparison of the DTT parameters, between the affected and unaffected hemispheres, for all the temporal projections showed significant differences in at least one DTT parameter (*p*<0.05). Only the UFZ showed significant differences in all the DTT parameters (*p*<0.05). The FA and FN ratios of the UF were strongly and positively correlated with the MMSE performed within one month of onset. In contrast, no significant correlation was observed between the MMSE within one month of onset and the DTT parameters of the cingulum, SLF, and ILF (*p*>0.05). The FA and FN ratios of the UF showed strong and positive correlations with the MMSE performed six months after onset, and the ADC ratio of the UF presented moderate and negative correlations (*p*<0.05). However, there were no significant correlations between the MMSE performed six months after onset and all the DTT parameters of the cingulum, SLF, and ILF (*p*>0.05).

**Table. 1.** Demographic data of the patients.

| Variables | Patients |
| --- | --- |
| Sex (male:female) | 17:8 |
| Mean age, year | 51.9(14.2) |
| Lesion side (right:left) | 14:11 |
| Duration from onset (months) | 16.9(18.0) |
| Mini-Mental State Examination at the same time as DTI scanning (within one month from onset) | 23.2(3.9) |
| Mini-Mental State Examination after six months from onset | 26.0(3.9) |

Values represent mean ( ± standard deviation), DTI: Diffusion tensor imaging.

**Table 2.** Results of the diffusion tensor tractography parameters for the temporal projections.

| Temporal projections |  | FA | ADC | FN |  |  |  |  |  |  |  |  |
| --- | --- | --- | --- | --- | --- | --- | --- | --- | --- | --- | --- | --- |
| Cingulum | Affected | 0.46 (0.05) | 0.83 (0.06) | 1051.7 (580.5) |  |  |  |  |  |  |  |  |
|  | Unaffected | 0.46 (0.04) | 0.80 (0.03) | 1222.8 (434.3) |  |  |  |  |  |  |  |  |
|  | p-value | 0.716 | 0.03* | 0.244 |  |  |  |  |  |  |  |  |
| SLF | Affected | 0.41 (0.04) | 0.8 (0.04) | 2613.4 (1498.1) |  |  |  |  |  |  |  |  |
|  | Unaffected | 0.44 (0.02) | 0.78 (0.03) | 2919.5 (1005.2) |  |  |  |  |  |  |  |  |
|  | p-value | 0.03* | 0.019* | 0.401 |  |  |  |  |  |  |  |  |
| ILF | Affected | 0.41 (0.04) | 0.88 (0.05) | 1704.1 (834.8) |  |  |  |  |  |  |  |  |
|  | Unaffected | 0.44 (0.02) | 0.85 (0.06) | 2229.4 (688.8) |  |  |  |  |  |  |  |  |
|  | p-value | 0.000* | 0.133 | 0.019* |  |  |  |  |  |  |  |  |
| UF | Affected | 0.37 (0.06) | 0.88 (0.15) | 363.5 (313.6) |  |  |  |  |  |  |  |  |
|  | Unaffected | 0.43 (0.02) | 0.81 (0.04) | 975.1 (513.3) |  |  |  |  |  |  |  |  |
|  | p-value | 0.000* | 0.024* | 0.000* |  |  |  |  |  |  |  |  |
| Correlation between the MMSE and the DTT parameters for temporal projections |  |  |  |  |  |  |  |  |  |  |  |  |
| MMSE | Cingulum |  |  | SLF |  |  | ILF |  |  | UF |  |  |
|  | FA ratio | ADC ratio | FN ratio | FA ratio | ADC ratio | FN ratio | FA ratio | ADC ratio | FN ratio | FA ratio | ADC ratio | FN ratio |
| One-month | -0.066 | -0.167 | -0.05 | 0.376 | -0.328 | 0.221 | 0.071 | -0.174 | -0.352 | 0.582* | -0.299 | 0.565* |
| Six-month | 0.260 | -0.104 | 0.166 | 0.217 | -0.003 | 0.156 | 0.187 | 0.034 | -0.252 | 0.525* | -0.424* | 0.580* |

Values represent mean ( ± standard deviation), FA: fractional anisotropy, ADC: apparent diffusion coefficient, FN: fiber number, SLF: superior longitudinal fasciculus, ILF: inferior longitudinal fasciculus, UF: uncinated fasciculus, MMSE: Mini-Mental State Examination, DTT: Diffusion tensor tractography.
\* $p < .05$

The result of the multiple regression analysis is shown in Table 3. The FN of the UF on the MMSE at six months from the onset had an explanatory factor of 33% and showed the 0.064 for unstandardized coefficients.

**Table 3.**
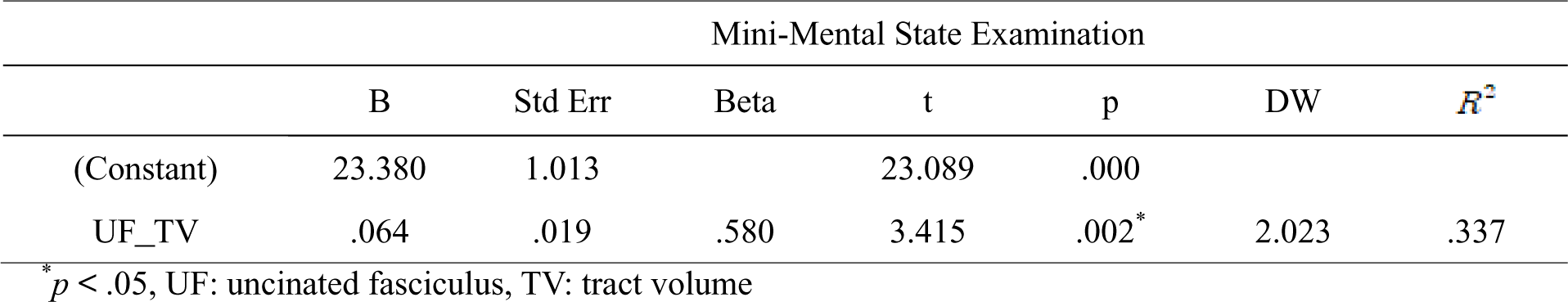
Results of a multiple regression analysis of the Mini-Mental State Examination.

## Discussion

In this study, the relationship between the early-stage temporal projections and the cognitive function at the chronic stage was investigated using DTT in patients with PH. Our results can be summarized as follows: (1) Of the DTT parameters of all temporal projections in the affected hemisphere, at least one parameter was affected compared to those of the unaffected hemisphere. These results indicated that the PH could possibly cause damage to all temporal projections regardless of whether the injury significantly affected cognitive function. (2) However, only the DTT parameters of the UF correlated with the MMSE performed within one month and after six months from onset. (3) The results of the multiple regression analysis revealed that the FN of the UF on the MMSE at six month had an explanatory power of 33% and showed the 0.064 for unstandardized coefficients. Hence, we believe that the UF would influence the cognitive function more than the other temporal projections. In particular, the early-stage FN of the UF could be an indicator of cognitive function at the chronic stage following PH. Among the DTT parameters, the FA, ADC, and FN have been widely used to detect injuries to the neural tracts.^22^ The FA value, ranging from 0 to 1, measures degree of anisotropy of water diffusion in the brain’s white matter and indicates the amount of myelination of the axons.^22^ The ADC value quantitatively expresses the degree of diffusion of water molecules through different tissues.^22^ The FN value indicates the volume of the neural tract.^22^ Therefore, a decreased FA or FN, or increased ADC, represents injuries to the neural tract. In detail, because the FN of the neural tracts can be visualized and directly reflects the state of the neural tract, many studies have reported that a decreased FN could be used as an indicator of injury to the neural tracts even without significant decreases in the FA or increases in the ADC.^33–35^ We believe that among the DTT parameters of the four temporal projections, the FN of the UF was the most significant factor influencing cognitive impairment following PH.

Over the last decade, various studies have reported changes in the DTT parameters of the temporal projections in patients with cognitive impairment following various brain injuries.^12,16,17,23–25^ In 2009, Kuniaki et al. demonstrated that significant differences in the FA and ADC values of the UF were observed between 17 patients with Alzheimer’s disease and 16 controls.^23^ In addition, they also found that a decreased FA value was observed in the posterior cingulum of 16 patients with mild cognitive impairment when compared with the 16 controls.^23^ In the following year, Morikawa et al. reported that the FA and ADC values of the UF were correlated with the MMSE in 30 patients with Alzheimer’s disease.^16^ In 2016, Singh et al. demonstrated that a decreased FA value of the UF showed a correlation with attention, spatial memory, sensorimotor dexterity, and emotion in 14 patients with schizophrenia ^17^. In the following year, Koyama and Domen investigated the relationship between injuries to the SLF and cognitive function in 40 patients with cerebral infarcts.^24^ The study suggested that the FA value of the SLF may be useful in predicting cognitive function after a cerebral infarct.^24^ In 2020, Chen et al. reported that the right inferior fronto-occipital fasciculus and the ILF could be markers for predicting cognitive dysfunction based on a study in 23 patients with white matter hyperintensity.^25^ The above studies indicated that the cingulum, SLF, ILF, and UF were associated with cognitive function and these projections could be possibly used as an indicator of cognitive function.^16,17,23–25^ Our results also suggest that though the DTT parameters indicated injuries to the cingulum, SLF, ILF, and UF in the affected hemisphere compared to those of the unaffected hemisphere, only the DTT parameters of UF showed a correlation with the MMSE. We believe that this result can be ascribed to the location of the PH which was mainly involved in the pathway of the UF.

Two DTT-based studies in PH patients have been reported earlier.^4,18^ Kwon et al. (2014) demonstrated that injuries to the anterior portion of the cingulum seen in 46% of 63 patients with PH affected the MMSE.^4^ In other words, this study also indicated that 54% of 63 patients who scored an average of 26.2 points on the MMSE did not have injuries of the cingulum.^4^ They just insisted that the anterior cingulum is vulnerable to PH. In 2018, Yang et al. investigated the fornix and cingulum in 16 chronic patients with left PH and found that only the left FN of the fornix was correlated with the intelligence quotient and the MMSE.^18^ In addition, they did not observe that the cingulum was not injured and not correlated with cognitive function.^18^ Thus, the two studies detailed above suggest that in PH, the cingulum might not be injured and may not be mainly associated with the cognitive function following PH.^4,18^ These results were in accordance with our results, which indicated that the cingulum was not correlated with the MMSE. In addition, our results showed that among the four temporal projections, the UF was the most significant predictor of the relationship between the temporal projections at an early stage and the cognitive function at the chronic stage. Therefore, we believe that the state of the UF could be used to predict the cognitive function following PH. However, our study has some limitations as follows: As this study was conducted retrospectively, we were not able to get detailed neuropsychological data, except for the MMSE. Hence, we could not investigate specific cognitive functions related to each temporal projection. In addition, both false-positive and negative DTT results could be presented due to the crossing fiber or partial volume effects.^36^

In conclusion, using DTT, we demonstrated that among the four temporal projections, the early state of the UF in patients with PH could predict long-term cognitive function. Our results suggest that the evaluation of the UF using DTT could be useful for predicting the cognitive function following PH.

**Figure 1.**
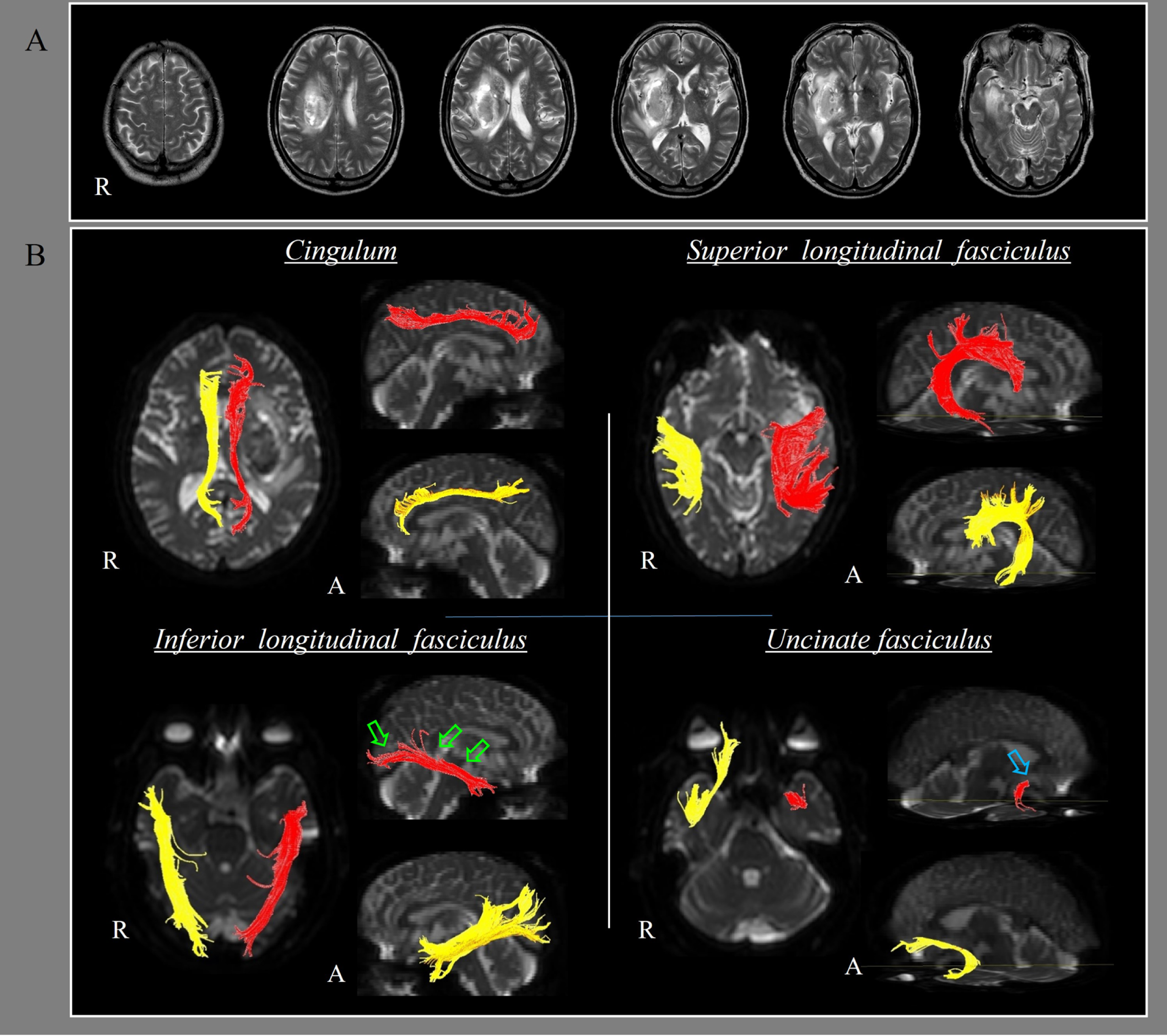
Results of the brain magnetic resonance imaging (MRI) and diffusion tensor tractography of the four temporal projections. (A) Brain MRI shows the putaminal hemorrhage. (B) Results of diffusion tensor tractography of the four temporal projections in a patient. Thinning (green arrows) of the left inferior longitudinal fasciculus and discontinuation (sky-blue arrow) of the left uncinated fasciculus were seen in the patient.

## Data Availability

The datasets generated during and/or analyzed during the current study are available from the corresponding author on reasonable request

